# *LTA4H* association with montelukast response in early and late-onset asthma

**DOI:** 10.1101/2020.10.05.20206847

**Authors:** Cyrielle Maroteau, Antonio Espuela-Ortiz, Esther Herrera-Luis, Sundararajan Srinivasan, Fiona Carr, Roger Tavendale, Karen Wilson, Natalia Hernandez-Pacheco, James D. Chalmers, Steve Turner, Somnath Mukhopadhyay, Anke-Hilse Maitland-van der Zee, Esteban G. Burchard, Maria Pino-Yanes, Simon Young, Glenda Lassi, Adam Platt, Colin N. A. Palmer, on behalf of the PiCA consortium

## Abstract

Leukotrienes play a central pathophysiological role in both pediatric and adult asthma. However, 35% to 78% of asthmatics do not respond to leukotriene inhibitors. To test the role of the *LTA4H* regulatory variant rs2660845 and age of asthma onset in response to montelukast in ethnically diverse populations.

We identified and genotyped 3,594 asthma patients treated with montelukast (2,514 late-onset and 1,080 early-onset) from seven cohorts (UKBiobank, GoSHARE, BREATHE, Tayside RCT, PAGES, GALA II and SAGE). Individuals under montelukast treatment experiencing at least one exacerbation in a 12-month period were compared against individuals with no exacerbation, using logistic regression for each cohort and meta-analysis.

While no significant association was found with European late-onset subjects, a meta-analysis of 523 early-onset individuals from European ancestry demonstrated the risk of experiencing asthma exacerbations in the G allele carriers’ group (AG or GG), despite montelukast treatment, was increased (odds-ratio=3.27, 95%confidence interval: 0.98–10.93, I2=69%, p=0.05) compared to those in the AA group. When meta-analyzing with other ethnic groups, no significant increased risk of asthma exacerbations was found (OR=1.69, 95% CI: 0.56-5.09, I2=84.81%, p=0.35).

Our study demonstrates that genetic variation in *LTA4H*, together with timing of asthma onset, may contribute to variability in montelukast response. Europeans individuals with early-onset (≤18y) carrying the rs2660845 G allele have increased risk of exacerbation under montelukast treatment, presumably due to the up-regulation of *LTA4H* activity. These findings support a precision medicine approach for the treatment of asthma with montelukast.

## INTRODUCTION

Asthma is a common chronic inflammation of the airways manifesting as shortness of breath, wheeze and chest tightness that can lead to death, especially in older adults. With around 339 million affected people worldwide (1) and an increase in incidence over recent decades, asthma is still under-diagnosed and under treated. The wide variation observed within asthma-related traits, including age of onset, respiratory symptoms, risk factors, frequency of exacerbations, lung function, comorbidities, and underlying inflammatory patterns, provide difficulty for diagnosis and are believed to contribute to heterogeneous treatment response. Precision medicine, by identifying the patient most likely to benefit from a particular therapy, may improve asthma control and reduce future risks, such as exacerbations and lung function decline.

Currently, biomarkers of airway inflammation, such as eosinophil count, immunoglobulin E (IgE), or fraction of exhaled nitric oxide (FeNO), are used clinically to target treatment and predict future risk (2). Disease severity, exacerbation triggers and comorbidities are also used as clinical biomarkers of asthma phenotype, with the age of onset of asthma and genetic risk factors recently emerging (3-7).

Leukotriene modifiers are pharmacological therapies for the treatment of asthma and allergic rhinitis. As established through numerous clinical trials, drugs such as montelukast have been shown to be effective in improving lung function, asthma control and quality of life, by reducing airway hyperresponsiveness and eosinophilia (8). Montelukast targets the cysteinyl leukotriene receptor type-1 (CysLTR1) at the cell membrane to block binding of cysteinyl leukotriene, thereby reducing CysLT-mediated airway bronchoconstriction [8]. Recent guidelines for the management of asthma in children and adults from the National Institute for Health and Clinical Excellence (NICE), highlighted leukotriene receptor antagonists (LTRA) as a key part of asthma management and recommended their use as the first line add-on therapy for persistent asthmatic patients who still show symptoms despite low dose of inhaled corticosteroids (ICS) (9). However, a range of 35 to 78 percent of the patients treated with leukotriene inhibitors such as montelukast were found unresponsive over independent studies (10-14).

Pharmacogenetic studies have investigated candidate genes in the leukotriene pathway, showing that interpatient variability might be influenced by genetic variation. Variants in *ALOX5, LTA4H, LTC4S, ABCC1, CYSLTR2*, and *SLCO2B1* genes may contribute to the heterogeneity of response to leukotriene modifiers, including montelukast (11, 15-17). To date, only the effect of a small number of genetic variants have been validated in independent studies (10, 18). We focused our analysis on a particular variant in the regulatory region of the leukotriene-A_4_ hydrolase (EC3.3.2.6) gene (*LTA4H*), the single nucleotide polymorphism (SNP) rs2660845. This variant has been shown to influence montelukast response through the prevention of exacerbations in a cohort of young adults with asthma from the United States (11). Association of pulmonary parameters such as peak expiratory flow (PEF) and forced expiratory volume in one second (FEV1) with rs2660845 has also been observed in a Japanese cohort [16]. Lima and colleagues highlighted a subgroup of responders to this drug class with the rs2660845 G allele as having four times the likelihood of experiencing an exacerbation compared to AA carriers (11) after six months of montelukast treatment. However, the sample size of this study was small, and the association observed between the rs2660845 SNP and responsiveness to montelukast has not been validated.

In this study, we utilized a very large dataset, consisting of previously described, ethnically diverse case-control studies (GALA II and SAGE), asthma cohorts (BREATHE and PAGES), a randomized clinical trial (RCT) (Tayside RCT) and two large real-life cohorts (the UKBiobank and GoSHARE, to explore the association of rs2660845 with lack of montelukast treatment response. We also wished to determine if age of asthma onset status further refined this putative pharmacogenomic association to provide the beginnings of a precision medicine strategy for the treatment of asthma with montelukast.

## METHODS

### Study design

Asthmatic patients treated with montelukast for at least 6 months were included in this study. We defined early-onset as individuals ≤18 years old at asthma diagnosis and late-onset for individuals over 18 years old at asthma diagnosis.

Early-onset individuals were recruited from five cross-sectional studies (PAGES, BREATHE, Tayside RCT, GALA II and SAGE) as well as from one longitudinal study (GoSHARE, the Genetics of Scottish Health Registry (19)). BREATHE, Tayside RCT and PAGES’ patients were recruited from primary and secondary care in Scotland. BREATHE and Tayside RCT details of enrollment have been presented in detail previously (20, 21). PAGES, the Pediatric Asthma Gene Environment Study (http://www.asthma-pages.com/), was an exploratory study of the genetic variation and exposure in asthmatic Scottish children from various ethnicities (African, Chinese, Indian, Pakistani, White British (93%), Mixed and other). Details of recruitment have been published earlier (22). The Genes-environments and Admixture in Hispanics/Latinos (GALA II) study and the Study of African-Americans, Asthma, Genes, and Environments (SAGE) recruited children from community- and clinic-based centers, with GALA II focused on Hispanics/Latinos and SAGE focused on African Americans (23). BREATHE, PAGES, GALA II and SAGE studies were all part of the Pharmacogenetics in Childhood Asthma (PiCA) consortium (24). For all studies from the PiCA consortium and Tayside RCT, asthmatic individuals under montelukast treatment were selected by a physician and information such as age, sex, BMI and OCS use, hospitalization and exacerbation were available from questionnaire filled at patient enrollment. In GoSHARE, asthma patients were selected from a dataset containing complete electronic medical records (EMR), prescription information, hospital and emergency room records (International Classification of Diseases 10th revision [ICD-10] codes J45) from Tayside, Scotland (Table E2).

Late-onset individuals were recruited from the UKBiobank (25) and GoSHARE. In the UKBiobank, selected asthma patients were British white individuals from self-reported questionnaires and hospital records (ICD10: J45) released in October 2019 by querying an AstraZeneca in-house protected UKBiobank database server. Other information on UKBiobank individuals such as age of onset of asthma and sex were extracted from self-reported questionnaires whereas montelukast usage was extracted from the recent release of drug prescriptions (October 2019).

The studies included in this article comply with the Declaration of Helsinki, and locally appointed ethics committee have approved respective research protocols and informed consent has been obtained from the subjects. PAGES was approved by the Cornwall and Plymouth Research Ethics Committee (Plymouth, United Kingdom). GoSHARE, BREATHE and Tayside RCT was approved by the Tayside Committee on Medical Research Ethics (Dundee, United Kingdom). The UKBiobank was approved by the National Research Ethics Committee. The Human Research Protection Program Institutional Review Board of the University of California, San Francisco (San Francisco, United States) approved GALA II and SAGE (ethics approval numbers: 217802 and 210362, respectively).

Asthma treatment was categorized following British Thoracic Society/ Scottish Intercollegiate Guideline Network (BTS/SIGN) guidelines (2019, https://www.brit-thoracic.org.uk/quality-improvement/guidelines/asthma/): step 1 as required short-acting beta-agonists (SABA) but no preventer, step 2 SABA plus inhaled corticosteroids (ICS) monotherapy or SABA plus LTRA, step 3 SABA plus ICS plus LTRA, and step 4 SABA plus ICS plus LTRA plus long-acting beta agonists (LABA) or SABA plus ICS plus LABA. Only individuals on montelukast therapy were selected for further analysis. Both late-onset and early-onset patients were selected from the GoSHARE study. As age of asthma onset was not reported in the EMR for GoSHARE, hence we use age at first salbutamol, age at first Inhaled corticosteroid and age at first montelukast prescription, all three over 18 years old to characterize the late-onset group GoSHARE (a) and all three under 18 years old to characterize the early-onset group GoSHARE (b).

### Binary risk of having an asthma exacerbation

The association between the variant of interest and the risk of having an asthma exacerbation was tested. A binary variable related to the absence or presence of at least one exacerbation event in a time frame of 6 to 12 months (0 = no exacerbation, 1 = at least one exacerbation event) was used. Asthma exacerbations were defined as below.

For PAGES, BREATHE and Tayside RCT, the definition of exacerbation was at least one of the following in the previous six months of recruitment: hospital admission, short course of oral corticosteroids (OCS) or absence from school, all due to asthma symptoms and verified by a general practitioner or a nurse.

For the UKBiobank and GoSHARE (a and b), the definition of exacerbation was at least one of the following within a year after first montelukast prescription date: hospital admission for asthma symptoms (ICD10: J45), emergency room visit (ER) for asthma symptoms (ICD10: J45), and two or more prescriptions of OCS.

For GALA II and SAGE, asthma exacerbations were defined by the presence of at least one of the following events in the 12 months preceding the study inclusion: need to seek emergency asthma care, hospitalization, or the administration of OCS due to asthma symptoms.

### DNA collection, extraction and analysis

For BREATHE and Tayside RCT, saliva was collected in commercially available kits (Oragene, DNA Genotech Ontario, Canada), DNA was prepared using the Qiagen DNeasy 96 kit (Qiagen, Manchester, UK) and genotypes were determined in the University of Dundee laboratory using TaqMan™ based allelic discrimination arrays on an ABI 7700 sequence detection system (ThermoFisher, Waltham, USA). For GoSHARE (a and b) respectively, DNA was extracted from blood samples taken at recruitment or from a prescribed blood test and genotyped using the Infinium Global Screening Array v2 (Illumina, San Diego, USA). Under the UKBiobank project, all recruited individuals were genotyped using a purpose-designed genotypic array: the UKBiobank Axiom Array. As part of the PiCA consortium, PAGES DNA was selected and genotyped in the Spanish National Genotyping Center (CEGEN-PRB3-ISCIII), using the Axiom Precision Medicine Research Array (PMRA) (Affymetrix, Thermo Fisher Scientific Inc, Waltham, MA). GALA II and SAGE were genotyped using the Axiom LAT1 array (World Array 4, Affymetrix, Santa Clara, CA, United States), as described elsewhere (26). Studies with genome-wide genotyping data were subjected to standard quality control procedures, ensuring that SNP genotyping call rate was above 95% and the absence of deviations from Hardy-Weinberg equilibrium (p>10-6) within control subjects. Samples with discrepancy between genetic sex and reported sex and with family relatedness were removed from the analyses.

### Statistical analysis

Statistical analysis of PAGES, BREATHE, Tayside RCT, the UKBiobank and GoSHARE (a and b) data were performed in SAS 9.3 (SAS Institute, Cary, NC, USA (27)) and PLINK 1.9 (28) for GALA II and SAGE. Binary logistic regression models were used to test the association between rs2660845 and asthma exacerbation status. Information related to age, gender, body mass index (BMI), asthma, age onset and exacerbation status a year before the start of therapy as well as the first two principal components of the genotype matrix, to control for population stratification, were included as covariates where appropriate and available for each study (Table E3). The effect of the *LTA4H* variant was considered dominant and, based on previous literature, the homozygous group for the major allele (AA) was the reference (see supplementary “Use of *LTA4H* rs2660845 variant as dominant”). Statistically significant associations were considered for p-values (P) lower than 0.05. Regarding the number of individuals selected we were sufficiently powered (>80%) to detect an OR of 1.5 or above based on our power calculation (Table E1) (29).

### Meta-analysis

Association results obtained from late-onset and early-onset studies were meta-analyzed separately to investigate the age of onset effect. For early-onset, populations of European origin were meta-analyzed together as well as with Latin/Hispanic and African American populations. Overall pooled odds ratios from all early-onset patients from GoSHARE(b), BREATHE, Tayside RCT, PAGES, SAGE, and GALA II studies, together with 95% confidence interval of the association between rs2660845 genotype and asthma exacerbation status, were obtained using a random effects model, because the phenotype definition as well as study characteristics varied among studies (Table 1). Whereas for late-onset Europeans studies (GoSHARE(a) and the UKBiobank), as the phenotype definition and baseline characteristics (age, gender, exacerbation percentage) were similar, a fixed effects model was used. The analysis was performed using the metafor package in R (30). We tested the heterogeneity among studies by means of the measure of inconstancy (I^2^) values as low (0-25%), moderate (25-50%) and high (50-75%). A threshold of P<0.05 was used to assess the statistical significance of the main effect association.

**Table 1.**
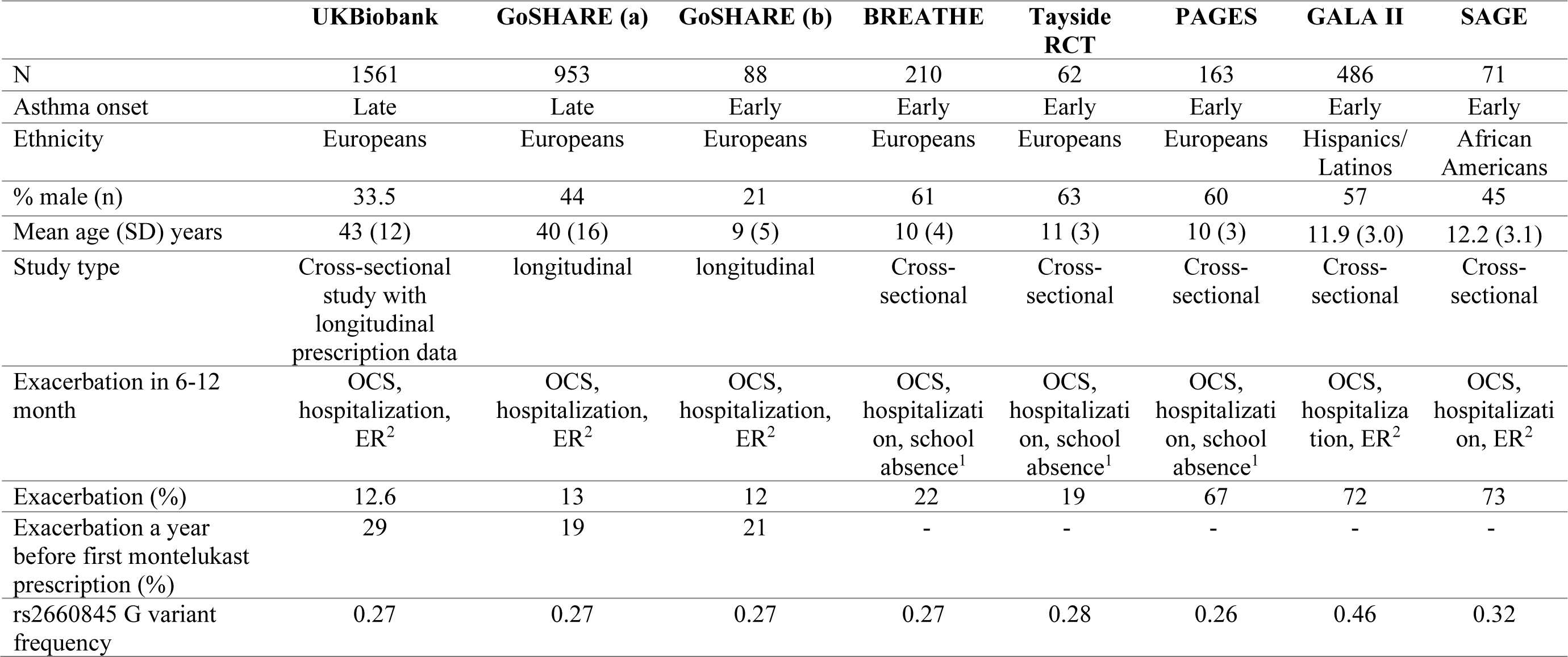
Characteristics of the studied cohorts. ^1^Exacerbation within 6 months; ^2^Exacerbation within 12 months. OCS: Oral Corticosteroids, ER: Emergency Room visit. Asthma onset is defined as early for individuals ≤ 18 years old at asthma diagnosis and late for individuals over 18 years old at asthma diagnosis.

All association tests compared those homozygous for the AA genotype with carriers of the G allele (AG or GG) (see supplementary “Use of LTA4H rs2660845 variant as dominant”).

## RESULTS

### Study characteristics

Study type and baseline characteristics of the participants for each study are presented in Table 1. A total of 3,594 individuals (2,514 late-onset and 1,080 early-onset) with asthma from multiple independent studies were used. More than 50% of the participants were female in SAGE, the UKBiobank and GoSHARE (a and b) (55%, 66.5%, 56% and 79% female respectively) and male in BREATHE, Tayside RCT, PAGES and GALA II (61%, 63%, 60%, 57% male respectively). The proportion of early-onset and late-onset individuals experiencing an exacerbation before being on montelukast was between 12% and 22% for BREATHE, Tayside RCT, GoSHARE and UKBiobank but was 67%, 72% and 73% respectively for PAGES, GALA II and SAGE. The minor allele frequency of rs2660845 in the UKB, GoSHARE, BREATHE, Tayside RCT, PAGES, GALA II and SAGE, study populations was 0.27, 0.27, 0.27, 0.28, 0.26, 0.46 and 0.32 respectively.

### No association between rs2660845 and exacerbation status in early or late-onset asthma in Europeans not under montelukast treatment

As a control analysis, we tested the association between rs2660845 and non-montelukast users in both late-onset and early-onset (see Supplementary materials for the selection of individuals and Table E4 for the cohort characteristics). No significant association was observed between rs2660845 and exacerbation rate in individuals early or late onset not under montelukast treatment (Table 2 and Table 3).

**Table 2.**
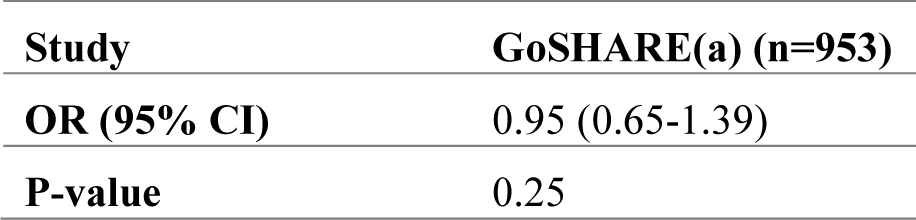
Association between rs2660845 and asthma exacerbation in late-onset GoSHARE(a) individuals a year before being on montelukast. Odds of exacerbation rate on montelukast for carriers of the minor allele (A/G or G/G) are being compared to homozygous for the ancestral allele (AA) in models adjusted for age, gender, age at 1st montelukast prescription or age at asthma onset and BMI when significant and available. GoSHARE(a): individuals with age at first salbutamol, age at first inhaled corticosteroid and age at first montelukast prescription, all three over 18 years old.

**Table 3.** Association between rs2660845 and exacerbation in individuals with early-onset asthma in PAGES and BREATHE non-montelukast users and in GoSHARE(b) individuals a year before being on montelukast. Odds of exacerbation rate on montelukast for carriers of the minor allele (A/G or G/G) are being compared to homozygous for the ancestral allele (AA) in models adjusted for age, gender, age at 1st montelukast prescription or age at asthma onset and BMI when significant and available. GoSHARE(b): individuals with age at first salbutamol, age at first inhaled corticosteroid and age at first montelukast prescription, all three under or at 18 years old.

| **Study** | **PAGES (n=356)** | **BREATHE (n=94)** | **GoSHARE(b)**<br>**(n=88)** |
| --- | --- | --- | --- |
| **OR (95% CI)** | 1.96 (0.92-4.18) | 0.93 (0.11-8.03) | 2.28 (0.91-5.67) |
| **P-value** | 0.17 | 0.94 | 0.43 |

### Association between rs2660845 and exacerbation status under montelukast treatment in Europeans with early-onset asthma

We assessed the risk of having an exacerbation whilst on montelukast treatment for carriers of the minor allele (AG or GG) compared to homozygous for the ancestral allele (AA). Table 4 shows the results of logistic regression models assessing the effect of rs2660845 (AG/GG against AA carriers) on exacerbation status. The genotypic effect was noted in three out of four European early-onset asthma studies. In BREATHE and Tayside RCT, individuals carrying at least a copy of rs2660845 G allele had respectively 4.4 (95%CI: 1.77-10.96, P=0.05) and 9.6 (95%CI:1.00-92.19, P=0.001) times the odds of having an exacerbation compared with AA genotype carriers, in a model adjusted for age at recruitment and BMI. In GoSHARE (b), the association was close to significance in a model adjusted for age at first LTRA prescribed and exacerbation a year before date of first montelukast prescription (Odds-ratio (OR) 9.8, 95% confidence interval (CI): 0.88-108.63; P=0.06). In PAGES the association was not significant (P=0.80).

**Table 4.** Association of exacerbation under montelukast treatment with rs2660845 genotype. Odds of exacerbation rate on montelukast for carriers of the minor allele (A/G or G/G) are being compared to homozygous for the ancestral allele (AA) in models adjusted for age, gender, age at 1st montelukast prescription or age at asthma onset and BMI when significant and available. Asthma onset is defined as early for individuals ≤ 18 years old at asthma diagnosis and late for individuals over 18 years old at asthma diagnosis.

| Odds ratio (95%CI) for exacerbation per G allele (referred to none); P |  |  |  |  |  |  |  |  |
| --- | --- | --- | --- | --- | --- | --- | --- | --- |
| Asthma onset | Late-onset |  | Early-onset |  |  |  |  |  |
| Ethnicity | Europeans | Europeans | Europeans | Europeans | Europeans | Europeans | Hispanics/<br>Latinos | African<br>Americans |
| Cohorts | UKBiobank | GoSHARE (a) | GoSHARE (b) | BREATHE | Tayside RCT | PAGES | GALA II | SAGE |
| **Montelukast**<br>**users** | 0.94 (0.69-1.27);<br>P=0.71 | 0.95 (0.64-1.41);<br>P=0.79 | 9.80 (0.88-108.63);<br>P=0.062 | 4.40 (1.77-10.96);<br>P=0.050 | 9.60 (1.00-92.19);<br>P=0.001 | 0.90 (0.41-1.95);<br>P=0.80 | 1.08 (0.68-1.71);<br>P=0.76 | 0.20 (0.05-0.84);<br>P=0.027 |

### Lack of an association between rs2660845 genotype and exacerbation status under montelukast treatment in Europeans late-onset asthma

No association between exacerbation risk and *LTA4H* rs2660845 was seen in any of the late-onset studies (Table 4).

### Association of rs2660845 with exacerbation status under montelukast treatment in Latino/Hispanic and African American early-onset asthma ethnic groups

In the Latino/Hispanic population, GALA II, the association was not significant (OR=1.08, 95%CI: 0.68-1.71, P=0.76). The African American population (SAGE) presented an opposite nominal association with an OR of 0.20 (95%CI: 0.05-0.83. P=0.027) in a model adjusted by age of asthma onset, gender and the first two principal components (Table 4).

### Meta-analyses of early-onset asthma cohorts

A meta-analysis of European early-onset cohorts PAGES, GoSHARE (b), BREATHE, Tayside RCT revealed that G allele carriers of rs2660845 SNP have 3.27 (95%CI: 0.98-10.93, P=0.05) times the odds of experiencing an exacerbation compared with patients with the AA genotype (Figure 1). When adding GALA II (Latino/Hispanic) and SAGE (African American), where rs2660845 MAF is higher than in European cohorts, no significant association between rs2660845 SNP and exacerbation status was found (OR=1.69, 95%CI: 0.56-5.10; P=0.35) (Figure 2).

**Figure 1.**
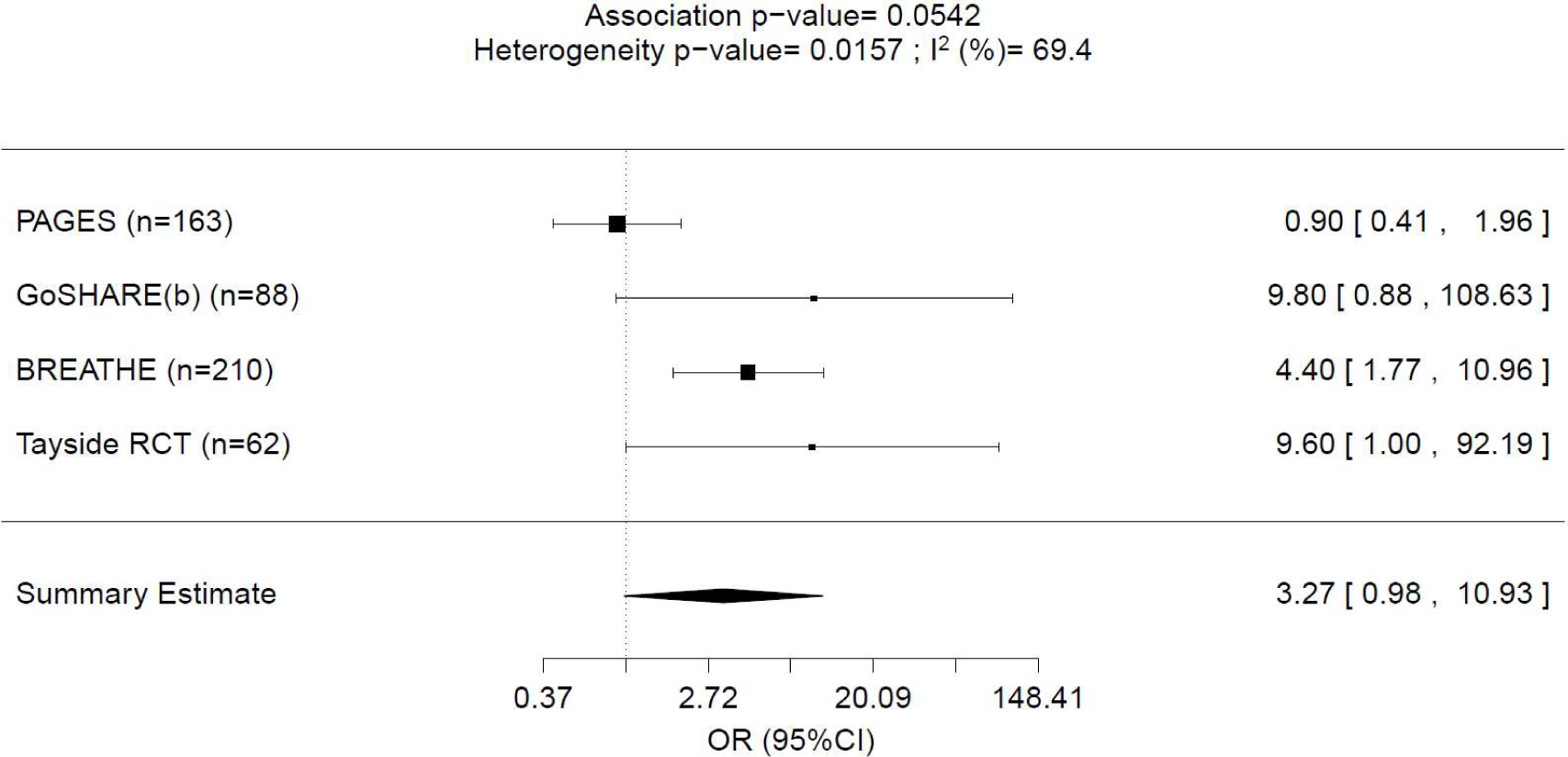
Forest plot representing meta-analysis of the association between *LTA4H* rs2660845 (AG+GG) and outcomes observed across early-onset PAGES, GoSHARE(b), BREATHE and Tayside RCT studies. Study sample size is in parentheses. Squares represent the odds ratio (OR), horizontal lines represent 95% confidence intervals (CIs). The diamond in the bottom represents summary estimate combining the study-specific estimates using a random effects model.

**Figure 2.**
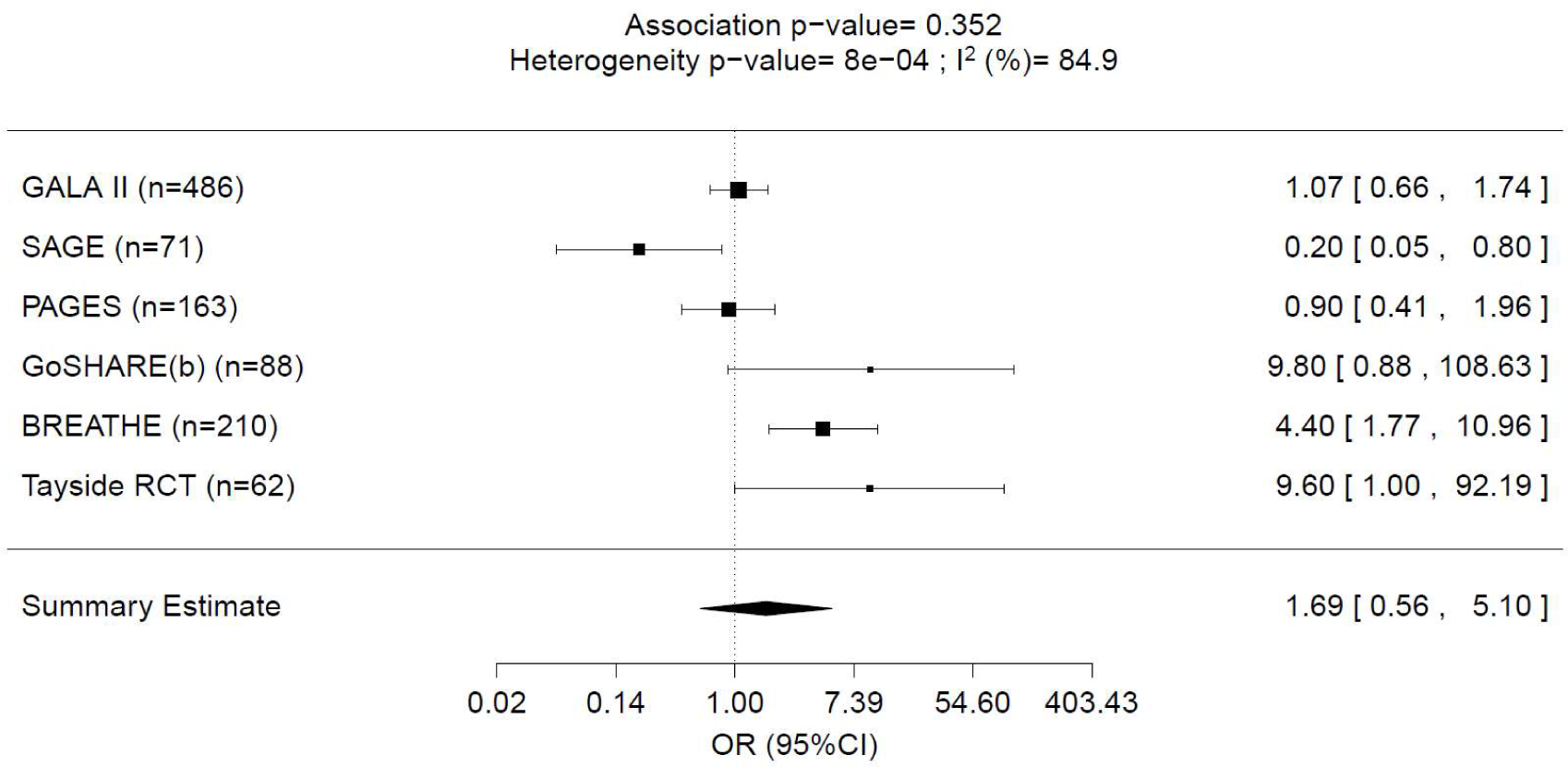
Forest plot representing meta-analysis of the association between *LTA4H* rs2660845 (AG+GG) and outcomes observed across all early-onset GALA II, SAGE, PAGES, GoSHARE(b), BREATHE and Tayside RCT studies. Study sample size is in parentheses. Squares represent the odds ratio (OR), horizontal lines represent 95% confidence intervals (CIs). The diamond in the bottom represents summary estimate combining the study-specific estimates using a random effects model.

### Meta-analysis of the European late-onset cohorts

A meta-analysis of the observed effects across late-onset populations of the UKBiobank and GoSHARE (a) showed no significant association between rs2660845 genotype and exacerbation status under montelukast treatment (OR= 0.94, 95%CI: 0.74-1.20; P=0.64) (Figure 3).

**Figure 3.**
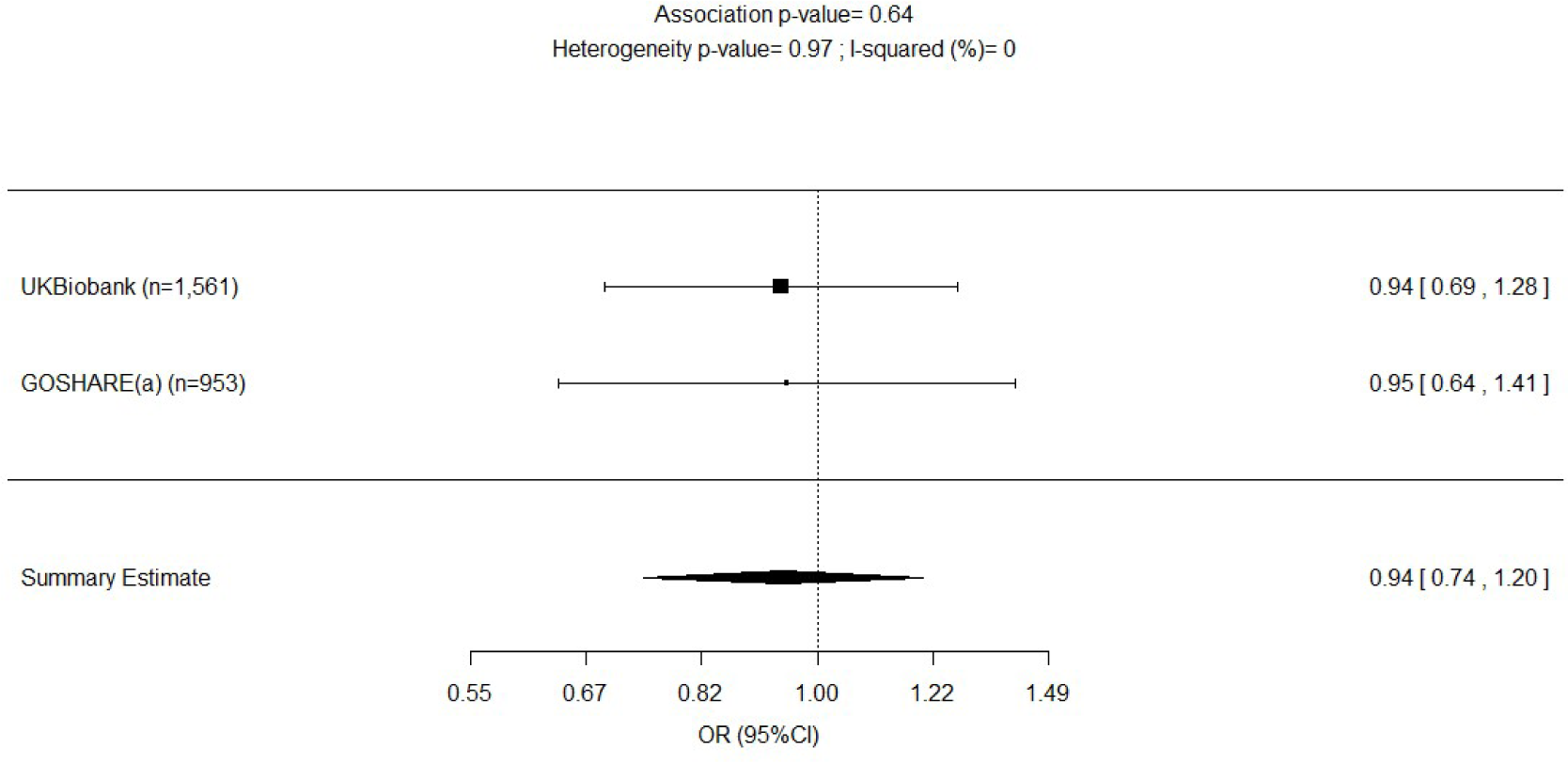
Forest plot representing meta-analysis of the association between *LTA4H* rs2660845 (AG+GG) and outcomes observed in late-onset UKBiobank and GoSHARE(a) studies. Study sample size is in parentheses. Squares represent the odds ratio (OR), horizontal lines represent 95% confidence intervals (CIs). The diamond in the bottom represents summary estimate combining the study-specific estimates using a fixed effects model.

## DISCUSSION

We report the results of the first study evaluating the association of the SNP rs2660845 with asthma exacerbations whilst on montelukast treatment, in late-onset and early-onset asthma patients, from different populations and ethnic backgrounds. We analyzed data from 3,594 individuals with asthma, 2,514 of which were late-onset and 1,080 early-onset.

Although no evidence of rs2660845 association with asthma exacerbations was found in late-onset cohorts of European ancestry, we observed a significant increased risk of exacerbation in early-onset asthma carriers of rs2660845 G allele with the European cohorts PAGES, GoSHARE(b), BREATHE and Tayside RCT (P= 0.05). Heterogeneous results were found in ethnically different early-onset populations for association between rs2660845 and risk for exacerbation whilst on montelukast treatment, with the risk being lower in the African American cohort SAGE and not significant in the Latino/Hispanic cohort GALA II. As our study identifies the rs2660845 genotype as associated with positive, negative and neutral effect on outcome in different populations, mechanistic studies would be required to confirm its relevance to LTRA response in different ethnic groups. However, rs2660845 may represent a robust biomarker for montelukast response in Europeans with early-onset asthma.

Since early-onset and late-onset asthma present fundamental differences both in the establishment of disease, as well as genetic variation and expression (3-7), we chose to analyze the effect of *LTA4H* rs2660845 SNP in each onset group. We found that the age of onset plays a role in the response to montelukast in European early-onset asthma patients, where the SNP rs2660845 was significantly associated with montelukast response as measured by exacerbation rate. These results were consistent with previous studies where early-onset asthma is presented as more genetic and allergic driven than late-onset which tends to be more environmental and lung centered (7, 31). Our analysis highlighted a significant role of the SNP rs2660845 in the response to montelukast in early-onset asthma of European ancestry, despite having less power to detect an effect compared to late-onset (n=523 vs 2,514) and the presence of heterogeneous exacerbation rates between European early-onset studies (PAGES presenting an exacerbation rate three to four times higher than the three other European early-onset cohorts). Like PAGES, GALAII and SAGE, presented a higher exacerbation percentage before montelukast treatment. It is possible that those this higher exacerbation percentage explains the high heterogeneity found between early-onset asthma cohorts and so the difference in the outcome. However, it was not possible to investigate these cohorts in depth to determine what was driving this difference. As environmental factors can modulate the effect of genetic variants upon asthma patients (32, 33) or drug response, and we were not able to investigate their impact on the exacerbation rate in PAGES, it is important to note that we might underestimate the effect of rs2660845 SNP on montelukast response in the European early-onset meta-analysis.

There are also significant disparities in asthma prevalence, mortality and drug response between ethnic groups (34). Here we reported different results from four European early-onset cohorts, one Latino/Hispanic and one African American, with rs2660845 MAF higher in GALA II and SAGE populations (results consistent with gnomAD (35)) compared to the European studies. Previous sequencing of *LTA4H* in GALA II revealed the prevalence of five polymorphisms (36) differing from those found in a European cohort by Holloway *et al* (37). It is possible that ancestry-specific difference in genetic architecture (linkage disequilibrium patterns) may explain why rs2660845 SNP shows no or significant opposite effects in different populations. Furthermore, heterogeneous effects may also be combined with differences in the environmental background between African American, Latino and European populations as well as insufficient sample size (32, 38).

An indirect role of rs2660845 on asthma exacerbation has been suggested by Rao *et al*., with the inactivation of *LTA4H* resulting in less exacerbations (39). Leukotriene-B_4_ (LTB_4_), the product of the degradation of leukotriene-A_4_ by LTA4H, plays a role in recruiting CD4^+^, CD8^+^ and T-cells and thus triggering lung inflammation and airway responsiveness (40). Therefore, an inactivation of LTA4H would affect those inflammatory cells recruitment by lowering LTB_4_ concentration in the blood. As LTB_4_ levels in the airways have been shown to increase in blood and bronchoalveolar lavage of asthma patients (41-43) and to correlate with asthma severity (44, 45), we examined mRNA expression data from whole blood in the GTEx (46), eQTLGen (47) and BIOSQTL (48) databases. We found that rs2660845 SNP is associated with *LTA4H* expression, with AA genotype lowering the expression (Figure E2 and Table E7). rs2660845 is present in the 5’UTR regulatory region of *LTA4H*, indicative of a putative cis-eQTL effect. However, the gene expression data analyzed was collected from adults of European ancestry (Figure E1 a and b for GTEx) and no significant association was found between rs2660845 and montelukast response in an adult cohort of early-onset asthma from the UKBiobank (cohort details available in table E5 and association results in table E6). As *LTA4H* expression and effects have been shown to be different in late-onset and early-onset asthma (7), it is possible that the results would be significant in a pediatric study. A possible mechanism in early-onset asthma is presented in Figure 4. By down regulating *LTA4H* activity, individuals with a rs2660845 AA genotype have a reduced LTB_4_ blood concentration resulting in less LTB_4_ driven exacerbations, simultaneously montelukast, acting as a cysteinyl receptor antagonist, will prevent other products of LTA_4_ degradation by leukotriene C4 synthase from binding to the receptor, thus reducing exacerbations through this arm of the LTA_4_ pathway. Therefore, LTA4H inhibitors, 5-Lipoxygenase Activating Protein inhibitors and Leukotriene-B_4_ receptor antagonists might represent a potential therapeutic strategy that could modulate key aspects of early-onset asthma (37, 49).

**Figure 4.**
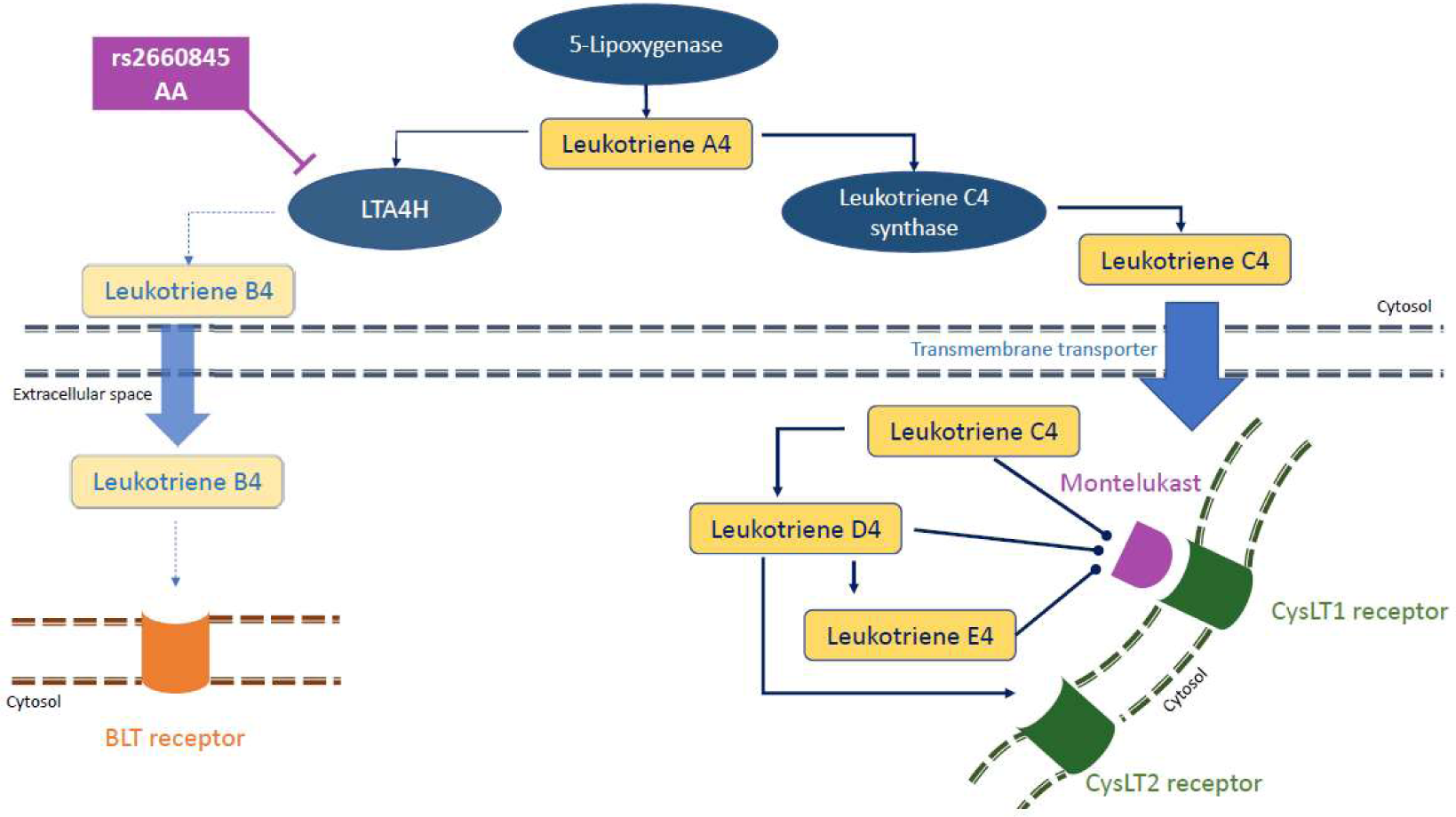
A possible mechanism of SNPs rs2660845 action on montelukast response in pediatric asthma patients. Reduced risk of exacerbation might be due to the down regulation of LTA4H activity, stimulating a montelukast sensitive pathway triggering exacerbation. rs2660845 AA genotype would result in a comparatively reduced LTA4H expression, leading to lowered LTB4 concentrations and so reducing exacerbation triggered by BLT receptors. Furthermore, LTA4 would be more readily converted into LTC4, unable to bind to CysLT1 receptors already blocked by montelukast.

Our study has limitations that should also be considered. A fine mapping of the *LTA4H* locus or haplotype analysis as reported by Holloway and colleagues (37) may help our understand the relationship between genetic variants of *LTA4H*, but was prevented by either GWAS or haplotype data being unavailable e for all cohorts. We decided to focus on rs2660845 as literature indicated a putative association with outcomes for asthma treatment (14). rs2660845 may be in linkage disequilibrium with yet undiscovered genetic variants that alter the gene expression or enzymatic activity so fine mapping and sequencing of the *LTA4H* region merits focused research in the future. Another important limitation is that we were not able to account for smoking, one of the most important environmental factors knowing its impact on lungs. Unfortunately, the information was not collected in a similar way for all studies, with only second-hand smoking information available for the pediatric cohorts (PAGES, BREATHE, Tayside RCT, GALAII and SAGE) and primary smoking status only available for a subset of GoSHARE and the UKBiobank cohorts. Finally, we have also assumed that treatment has been assigned based on similar prescribing criteria. However, we did not investigate other treatments and it is possible that individuals with increased risk of exacerbation might not have received LTRA treatment which would tend to underestimate the effect of the interaction between montelukast and rs2660845 for exacerbations. As other treatments were not taken into account, the effect found in the early-onset asthma group could also be overestimated and mostly linked to a more serious disease.

To our knowledge, this is the only study of leukotriene response to utilize populations of different age of onset and different ethnic backgrounds. These results, together with published studies in other European (37) and Latino/Hispanic cohorts (36), support a strong involvement of age of onset, LTA4H and LTB4 production in the pathophysiology of asthma. In conclusion, Europeans with early-onset asthma, carrying the G allele present in 27% of Europeans, and receiving montelukast were at 3.27 times increased risk for exacerbation compared to peer AA homozygotes. Our results add to the growing evidence of age of asthma onset as a robust clinical biomarker and moreover suggests stratifying treatment by age of asthma onset and rs2660845 status can form the basis of a precision medicine strategy to select those patients most likely to benefit from montelukast treatment.

## Data Availability

Data available upon request

## ACKNOWLEDGEMENTS

We would like to thank all the participants from the different cohorts, the BREATHE, Tayside RCT, PAGES, UKB, SHARE, GALA II, and SAGE teams as well as nurses, interviewers, technicians, clinicians and researchers’ volunteers and study coordinators, for their help recruiting the participants. CM would like to thank her colleagues from the Centre of Genomic Research (AstraZeneca) and specially Amanda O’Neil, Katherine Smith, Slavé Petrovski, Olympe Chazara, Sophia Cameron-Christie, Eleanor Wigmore, Niedzica Camacho, Dimitrios Vitsios and Sebastian Wasilewski for their help with the UKBiobank data. We acknowledge the support of the Health Informatics Centre, University of Dundee for managing and supplying the anonymized data.

