## Supplemental file for "*LTA4H* association with montelukast response in early and late-onset asthma"

**Contents**

Use of *LTA4H* rs2660845 variant as dominant

Association between rs2660845 and asthma exacerbation in non-montelukast users

Table E1

Table E2

Table E3

Table E4

Table E5

Table E6

Figure E1

Figure E2

References

**Use of *LTA4H* rs2660845 variant as dominant**

The association between the genotype and exacerbation status under montelukast treatment has been presented as an additive and dominant model in two other publication (respectively(E1, E2)). The sub-additivity of the effect is observed there. The use of a dominant model slightly underweights the effect of individuals homozygous for the minor allele; however, in order to perform a case-control analysis on small-sized cohorts we were under-powered to detect meaningful effects in the rare homozygotes. Moreover, the collapsing of the rare allele into a "carriers vs. non-carrier" group is common practice in order to reduce the volatility of small cell sizes in case control analyses.

**Association between rs2660845 and asthma exacerbation in non-montelukast users**

To verify the effect of rs2660845 on montelukast response and not just on exacerbation, we tested the association between rs2660845 and exacerbation in non-montelukast users. In late-onset, as we have longitudinal data for GoSHARE(a) cohort, individuals previously selected as being under montelukast treatment where used. The binary effect of experiencing an exacerbation was defined a year before start of therapy. In early-onset asthma, genetic data from non-montelukast users were only available from PAGES, BREATHE GALA and SAGE cohorts. Individuals were selected as not being under montelukast treatment at enrolment. The binary risk of having an asthma exacerbation was defined as 0 = no exacerbation, 1 = at least one exacerbation event, in a time frame of 6 to 12 months. An asthma exacerbation was defined as:

For PAGES and BREATHE, at least one of the following in the previous 6 months: hospital admission, course of oral corticosteroids (OCS) and absence from school due to asthma symptoms.

For GoSHARE (a), at least one of the following within a year before first montelukast prescription date: hospital admission, emergency room visit, course of OCS and discontinuation of the drug. Study details are presented in *Table E3*.

Statistical analyses were performed in SAS 9.3 (SAS Institute, Cary, NC, USA). Binary logistic regression model was used to test the association between rs2660845 and the binary risk of having an exacerbation. LTA4H variant effect was considered dominant and p value (P) less than 0.05 was considered significant. Results for late-onset and early-onset are presented in respectively *Table E4 and E5*.

**Table E1. Power of the sample size for combined cohorts to detect increases in OR for asthma exacerbation.**

GoSHARE (a) is the late-onset population (>18 years-old)

GoSHARE (b) is the early-onset population (<= 18 years-old)

| Asthma onset | Ethnicity | Cohort | OR=1.2 | OR=1.5 | OR=3 |
|---|---|---|---|---|---|
| Late | Europeans | UKBiobank, GoSHARE (a) | 82% | 99% | 100% |
| Early | Europeans | GoSHARE (b), BREATHE, Tayside RCT, PAGES | 31% | 86% | 100% |
| Early | Europeans, African Americans, Hispanics/Latinos | GoSHARE (b), BREATHE, Tayside RCT, PAGES, SAGE, GALA II | 64% | 97% | 100% |

**Table E2. Selection of asthmatic patients in GoSHARE by treatment steps.**

| Steps | Medication | Number of GoSHARE patients (n) |
|---|---|---|
| 1 | SABA as needed | 10,218 |
| 2 | SABA as needed +ICS | 5,388 |
| 3 | SABA as needed + ICS + LABA | 4,551 |
| 4 | SABA as needed + LTRA (+ICS +LABA) | 1,070 |

Step 1: inhaled short-acting β2-agonists (SABA) on demand;

Step 2: regular inhaled steroids (ICS) plus SABA on demand;

Step 3: regular inhaled long-acting β2-agonists (LABA) (salmeterol or formoterol) plus ICS with SABA on demand;

Step 4: oral montelukast with SABA on demand (plus ICS plus/or regular LABA).

**Table E3. Covariates association with asthma exacerbation binary trait.**

*Traits with a P <0.05 were used as covariates in the logistic regression.

For GALA II and SAGE, betas 95%(CI) are reported for quantitative variables

| | Odds ratio (95%CI) for exacerbation; P-value | | | | | | Betas (95%CI) for exacerbation; P-value | |
|---|---|---|---|---|---|---|---|---|
| | UKBiobank N=1,561 | GoSHARE (a) N=953 | GoSHARE (b) N=88 | BREATHE N=210 | Tayside RCT N=62 | PAGES N=163 | GALA II N=486 | SAGE N=71 |
| **Gender (M vs F)** | 1.07 (0.78-1.46) | 1.18 (0.76-1.82) | 12.9(2.10-79) | 1.04 (0.81-1.34) | 0.97 (0.31-2.71) | 1.99 (1.02-3.9); | 1.16 (0.75-1.81) | 2.39 (0.74-7.76) |
| | P=0.66 | P=0.44 | P=0.006* | P=0.74 | P=0.89 | P=0.04* | P=0.494 | P=0.1465 |
| **Age at 1st LTRA** | - | 1.01 (1.003-1.03) | 1.14(0.89-1.45) | - | - | - | - | - |
| | - | P=0.013* | P=0.29 | - | - | - | - | - |
| **Age at 1st SABA** | - | 1.01 (0.99-1.02) | 0.79 (0.56-1.09) | - | - | - | - | - |
| | - | P=0.18 | P=0.15 | - | - | - | - | - |
| **Exacerbation before 1st LTRA** | 1.4 (1.02-1.92) | 6.02 (3.91-9.26) | 8.91 (5.99-13.2) | - | - | - | - | - |
| | P<0.036* | P<0.0001* | P<0.0001* | - | - | - | - | - |
| **Age at recruitment** | - | - | - | 0.86 (0.83-0.89) | 1.11 (0.96-1.29) | 0.93 (0.84-1.02) | 0.03 (-0.01-0.07) | -0.07 ((-0.02) – (-0.18)) |
| | - | - | - | P<0.0001* | P=0.14 | P=0.16 | P=0.453 | P=0.476 |
| **BMI** | - | - | - | 0.95 (0.92-0.98) | 1.07 (0.94-1.22) | 0.97 (0.91-1.05) | - | - |
| | - | - | - | P=0.0035* | P=0.24 | P=0.54 | - | - |
| **Age of asthma onset** | 1.06 (1.02-1.09) | - | - | - | - | - | -0.07 [-0.10- (-0.03)] | -0.06 ((-0.25)-0.14) |
| | P=0.0003* | - | - | - | - | - | P=0.052* | P=0.5756 |
| **PC1** | - | - | - | - | - | - | 47.87 [-55.85-(-39.89)] | 19.24 ((-6.85)-45.32) |
| | - | - | - | - | - | - | P<0.0001* | P=0.148 |
| **PC2** | - | - | - | - | - | - | 5.28 (-2.14-12.71) | -0.60 ((-29.23)- 28.03) |
| | - | - | - | - | - | - | P=0.476 | P=0.967 |

**Table E4. Details of late-onset and early-onset in three non-montelukast user populations.**

[1]Exacerbation within 6 months; [2]Exacerbation within 12 months

OCS: Oral Corticosteroids

ER: Emergency Room visit

GoSHARE (a) is the late-onset population (>18 years-old)

GoSHARE (b) is the early-onset population (<= 18 years-old)

| | **GoSHARE (a)** | **GoSHARE (b)** | **BREATHE** | **PAGES** |
|---|---|---|---|---|
| N | 953 | 88 | 94 | 356 |
| % male (n) | 44 | 21 | 57 | 56 |
| Mean age (SD) years | 40 (16) | 9 (5) | 10 (3.6) | 10 (3.5) |
| Study type | longitudinal | longitudinal | Cross-sectional | Cross-sectional |
| Exacerbation in 6-12 month | OCS, hospitalisation, ER[2] | OCS, hospitalisation, ER[2] | OCS, hospitalisation, school absence[1] | OCS, hospitalisation, school absence[1] |
| Exacerbation (%) | 19 | 12 | 36 | 61 |
| rs2660845 G variant frequency | 0.27 | 0.27 | 0.27 | 0.26 |

**Table E5. Details of early-onset asthma adult montelukast user from the UKBiobank.**

[1]Exacerbation within 6 months; [2]Exacerbation within 12 months

OCS: Oral Corticosteroids

ER: Emergency Room visit

Patients were diagnosed as having early-onset asthma.

Montelukast prescription records were only available as adults.

| | UKBiobank |
|---|---|
| N | 511 |
| % male (n) | 34 |
| Mean age (SD) years | 10 (4) |
| Study type | longitudinal |
| Exacerbation in 6-12 month | OCS, hospitalisation, ER[2] |
| Exacerbation (%) | 21 |
| rs2660845 G variant frequency | 0.27 |

**Table E6. Association between rs2660845 and asthma exacerbation in early-onset UKBiobank individuals a year after taking montelukast prescription as adults.**

Patients were diagnosed as having early-onset asthma.

Montelukast prescription records were only available as adults.

| Study | UKBiobank (n=511) |
|---|---|
| **OR (95% CI)** | 1.34 (0.82-2.19) |
| **P-value** | 0.23 |

**Table E7. Cis-eQTL effect of rs2660845 on *LTA4H* expression in whole blood from adult cohorts.**

BIOSQTL: The Biobank-Based Integrative Omics Study Quantitative Trait Locus (E3)

eQTLGen: expression Quantitative Trait Loci Genetic (E4)

| Consortium | P-value | rsID | Chr | Position(hg19) | ID | Gene symbol | Z-score | Assessed | Other | Number of cohorts | Number of samples | FDR |
|---|---|---|---|---|---|---|---|---|---|---|---|---|
| **BIOSQTL** | 6.27E-08 | rs2660845 | 12 | 96438553 | ENSG00000111144,ENSG00000257878 | LTA4H | -5.41 | G | A | 4 | 2116 | 0 |
| **eQTLGen** | 4.89E-07 | rs2660845 | 12 | 96438553 | ENSG00000111144 | LTA4H | -5.03 | G | A | 37 | 31683 | 0.0016 |

FDR: False Discovery Rate

ID: Gencode Identifier

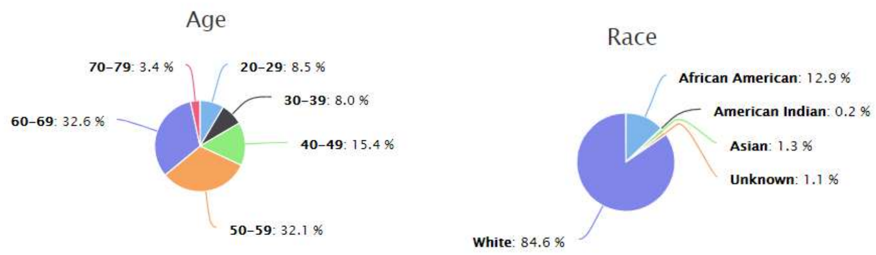


a b

**Figure E1**. **Age (a) and ethnicity (b) distributions in the GTEx portal (V8, (E5))**

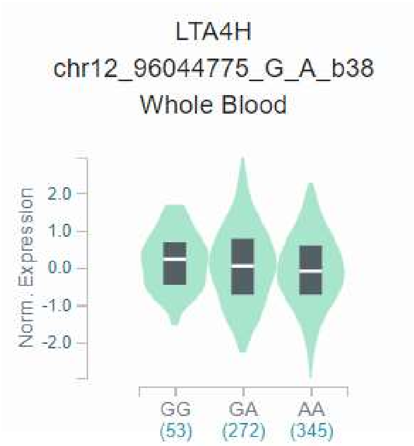


**Figure E2**. **Box plot showing the *cis*-eQTL effect of rs2660845 on $LTA_4H$ expression in whole blood (beta=-0.020, P-value=0.46). (GTEx v8, (E5))**
